# Validation of non-contact sensor quantification of heart rate and respiratory rate dynamics using real-world pretraining and label-efficient fine-tuning on polysomnograms

**DOI:** 10.64898/2026.07.01.26356016

**Authors:** Keshav S. Gupta, Nicholas Harrington, Raimon Padrós-Valls, Pamela DeYoung, Robert L. Owens, Jeremy E. Orr, Kevin R. King

## Abstract

Non-contact mechanical bed sensors can passively and longitudinally monitor the dynamics of cardiopulmonary physiology to detect changes from patient-specific baselines and facilitate care. This requires accurate longitudinal quantification of established metrics like respiratory rate (RR) and heart rate (HR) from the underlying raw waveforms, validated against ground truth labeled datasets like simultaneous polysomnography (PSG). Whereas head-to-head labeled datasets are scarce and costly to collect, unlabeled real-world datasets are often abundant. Here, we show that non-optimized heuristic algorithms can be used to “soft-label” large real-world data (*>*40M minutes across *>*50,000 nights) for pretraining models. This enables label-efficient fine-tuning on small numbers of head-to-head PSG-labeled datasets while maximizing generalizability and robustness to hyperparameters. The result is a highly performant validated model with mean absolute errors (MAE) of 0.6 brpm for RR and 1.1 bpm for HR across 1-minute windows. Although demonstrated in the context of bed sensor cardiopulmonary quantification, these methods are applicable to development of sensor algorithms whenever labeled ground truth data is scarce and unlabeled real-world data is abundant.

## Introduction

Continuous home physiological monitoring has the potential to improve patient outcomes by enabling early detection of clinical deterioration [1, 2]. To achieve this goal, diverse wearable and unobtrusive nearable sensors have been developed to dynamically monitor physiology and provide explainable outputs across time [3, 4]. Doing so often requires development of algorithms or machine learning models to accurately estimate established physiological metrics such as heart rate (HR) and respiratory rate (RR) from underlying raw sensor waveforms [5, 6]. Deep learning is particularly attractive because it can be trained to produce highly generalizable models that are robust to real-world variation, artifacts, and noise [7, 8]. However, they require training on labeled datasets, which can be challenging and costly to collect [9, 10]. Often, these sensors have abundant real-world data that cannot be used for training because it lacks labels from head to-head comparison with a validated sensor. Here we ask whether soft-labeling the abundant real-world data using a heuristic (i.e., “rule-based”) algorithm can be used to improve training efficiency on small, labeled datasets, (i.e., label-efficient fine-tuning), using non-contact bed sensors.

Non-contact bed sensors are attractive for longitudinal home monitoring because they maximize adherence and minimize missing data. Once placed beneath a mattress, these passive sensors can automatically transduce mechanical forces generated by each breath, heartbeat, and musculoskeletal movement during sleep without requiring charging, maintenance, or other user interaction [11]. One approach to quantification of the resulting data uses heuristic algorithms based on peak-finding and peak-counting [12]. These often perform well in controlled experimental settings but can be brittle when applied to real-world data. Deep learning models can be developed to predict vital sign metrics like HR and RR from raw sensor waveforms but doing so requires simultaneous collection with ground truth datasets, for example from head-to-head polysomnography (PSG) with electrocardiography for HR and respiratory inductance plethysmography for RR [13, 14, 15]. By comparison, real-world data is often abundant but is unlabeled.

Here we leverage large volumes of unlabeled real-world bed sensor data for pretraining cardiopulmonary quantification models by applying imperfect “soft labels” using non-optimized heuristic algorithms. This effectively bootstraps the model so that it can be fine-tuned on minimal head-to-head labeled data. This also exposes it to real-world diversity that can maximize generalizability of the final trained model. Our results show that this pretraining strategy results in a more sample-efficient, more hyperparameter-robust, and more noise-tolerant model when compared to randomly-initialized models. As the amount of available labeled data increases, the benefits of pretraining decline and are eventually lost. Together, our results suggest heuristic soft-labeling of real-world sensor data can be used to achieve label-efficient fine-tuning when head to-head labeled data is scarce. Although we demonstrate this in the context of bed sensor cardiopulmonary quantification, the method is applicable to development of sensor algorithms wherever labeled ground truth data is scarce and unlabeled real-world data is abundant.

## Results

### Collection of unlabeled and labeled sensor datasets

To test the hypothesis that large amounts of unlabeled real-world data can improve the efficiency of training on head-to-head data with ground truth labels, we leveraged two bed sensor datasets from Nightingale Labs (Fig. 1). Both were collected by a thin flexible ferroelectret mechanical sensor slipped under the mattress and plugged into the wall for indefinite adherence-independent monitoring. No restrictions were placed on mattress thickness, type, bed shape or size; sleeping position or sleep patterns; or number of sleepers. Dataset #1 was a large real-world unlabeled dataset comprising *∼*45 million minutes from 395 subjects collected across 50,848 nights of sleep. Dataset #2 was a focused 106-subject head-to-head labeled dataset in which the 100Hz bed sensor waveforms were collected during simultaneous professionally-annotated polysomnography, which included validated chest and abdominal Respiratory Inductance Plethysmography (RIP) and electrocardiography (ECG) from which RR and HR could be derived as ground truth labels. Our overall strategy used a heuristic peak-finding algorithm to apply “soft labels” to the large real-world Dataset #1, which enabled pretraining of a deep learning model on the soft-labeled data, and subsequent fine-tuning of the pretrained model using a subset of head-to-head PSG-labeled Dataset #2 followed by evaluation on a held-out segment of Dataset #2 (Fig. 1).

**Figure 1.**
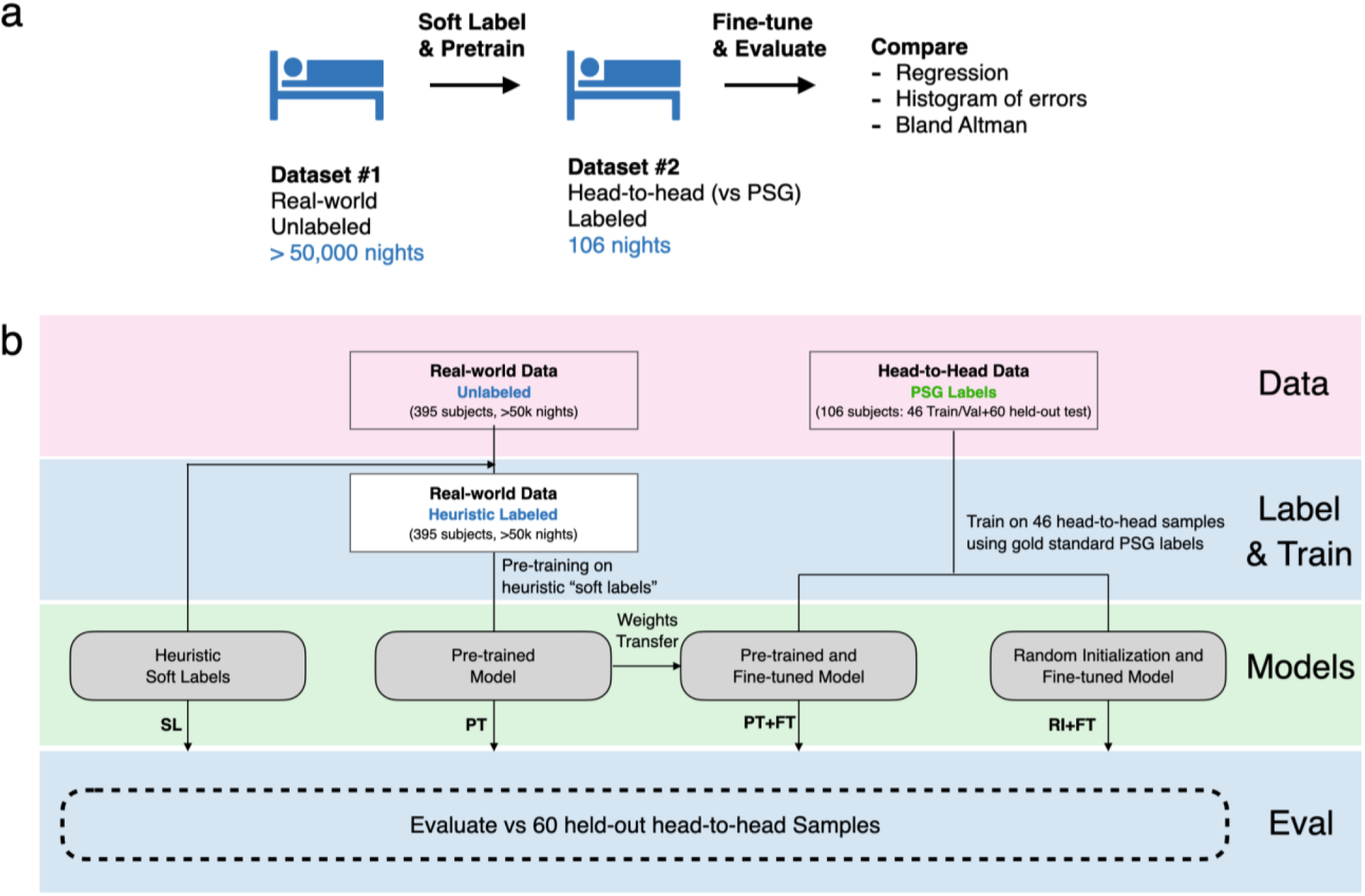
Soft-label pretraining and label-efficient fine-tuning of bed-sensor cardiopulmonary quantification models. (a) Unlabeled real-world bed-sensor recordings (Dataset #1) were soft-labeled with a heuristic algorithm and used to pretrain deep learning models, which were then fine-tuned on a smaller head-to-head PSG-labeled corpus (Dataset #2) and evaluated against PSG-derived ground truth. (b) Four sets of models were compared on the same 60-night held-out test set: the heuristic soft labels (SL), the pretrained model without fine-tuning (PT), the pretrained model fine-tuned on 46 PSG-labeled nights (PT+FT), and a randomly initialized model fine-tuned on the same 46 nights (RI+FT).

### Pretraining on real-world data using heuristic “soft labels”

We first focused on Dataset #1 and created pretrained models from unlabeled real-world data. Sensor waveforms were first separated into low and high frequency signals representing respiratory- and ballistocardiogram-predominant waveforms respectively. Next, we used peak-counting algorithms to estimate HR and RR soft labels at 1-minute windows from the underlying waveform. After applying task-specific quality filters (see methods for details) we were left with 15 million HR and 43 million RR epochs for training deep learning models (using an architecture comprised of a stack of one-dimensional convolutions feeding a bidirectional gated recurrent unit, whose output is projected by a multilayer perceptron to a scalar rate) to predict the heuristic soft-labels from the corresponding windows of Dataset #1 waveform.

### Fine-tuning and evaluation on held-out labeled datasets

We next performed fine-tuning using Dataset #2 (n=106 patients), the head-to-head bed sensor waveforms and labeled PSGs, which we divided into n=46 training and validation patients and a n=60 held-out test set of patients. To investigate the benefits of soft label pretraining, we compared the performance on the evaluation dataset with heuristic soft labels (SL), the pretrained model without fine-tuning (PT), the pretrained model with fine-tuning (PT+FT), and a randomly initialized model with the same fine-tuning (RI+FT) for their ability to predict HR and RR on the 60-night evaluation held-out test set (Fig. 1b, Tables 2–3). The combined pretrained and fine-tuned (PT+FT) models, when tested on the held-out dataset, achieved a mean absolute error (MAE) of 1.1 bpm for HR and 0.6 brpm for RR as demonstrated by regression, histogram of error, and Bland-Altman plots for HR and RR (Fig. 2). For HR, fine-tuning substantially improved performance compared to soft labels and the pretrained model alone. When trained using the maximal number of patients, fine-tuned models developed from pretrained initial weights were not substantially better than those starting with random initialization. For RR, the performance of the SL, PT, and PT+FT models were similar and each markedly outperformed RI+FT models suggesting that the initial heuristic algorithm and resulting soft labels were highly performant and models with access to these high-quality soft labels reproducibly outperformed those without (Fig. 3, Tables 2–3).

**Figure 2.**
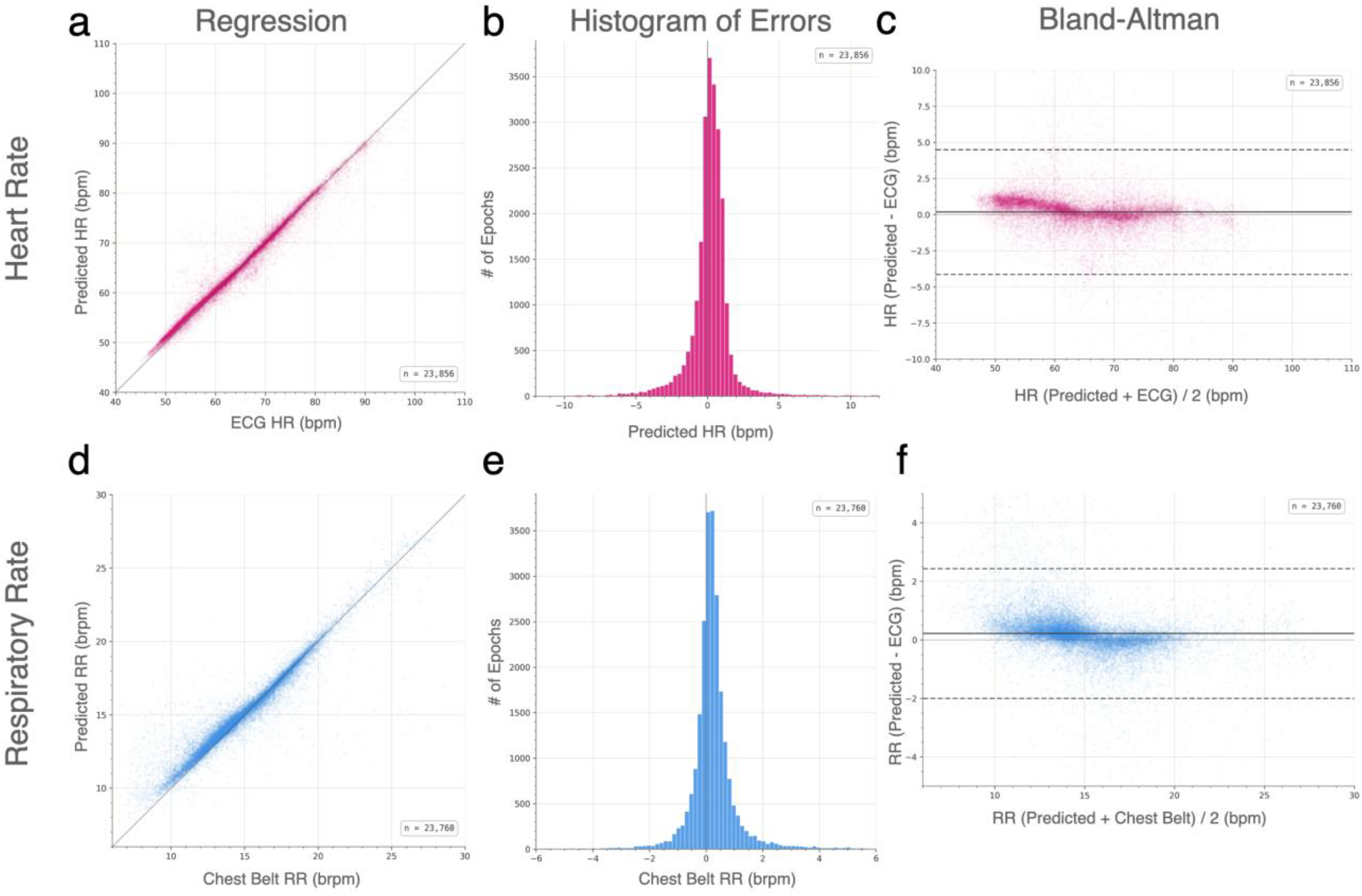
High performance bed sensor derived cardiopulmonary quantification enabled by pretraining on unlabeled data and label-efficient fine-tuning. Bed sensor derived Heart rate (HR) and respiratory rate (RR) were estimated from bed sensor waveforms at 1-minute intervals using models pretrained (PT) on unlabeled real-world data “soft labeled” with a heuristic algorithm followed by fine-tuning (FT) on n=46 PSG-labeled studies and evaluated on n=60 PSG-labeled held-out test data. Performance is shown for A-C) HR and D-F) RR models by A,D) regression (model prediction vs ground truth PSG labels), B,E) histogram of errors, and C,F) bland-altman plots.

**Figure 3.**
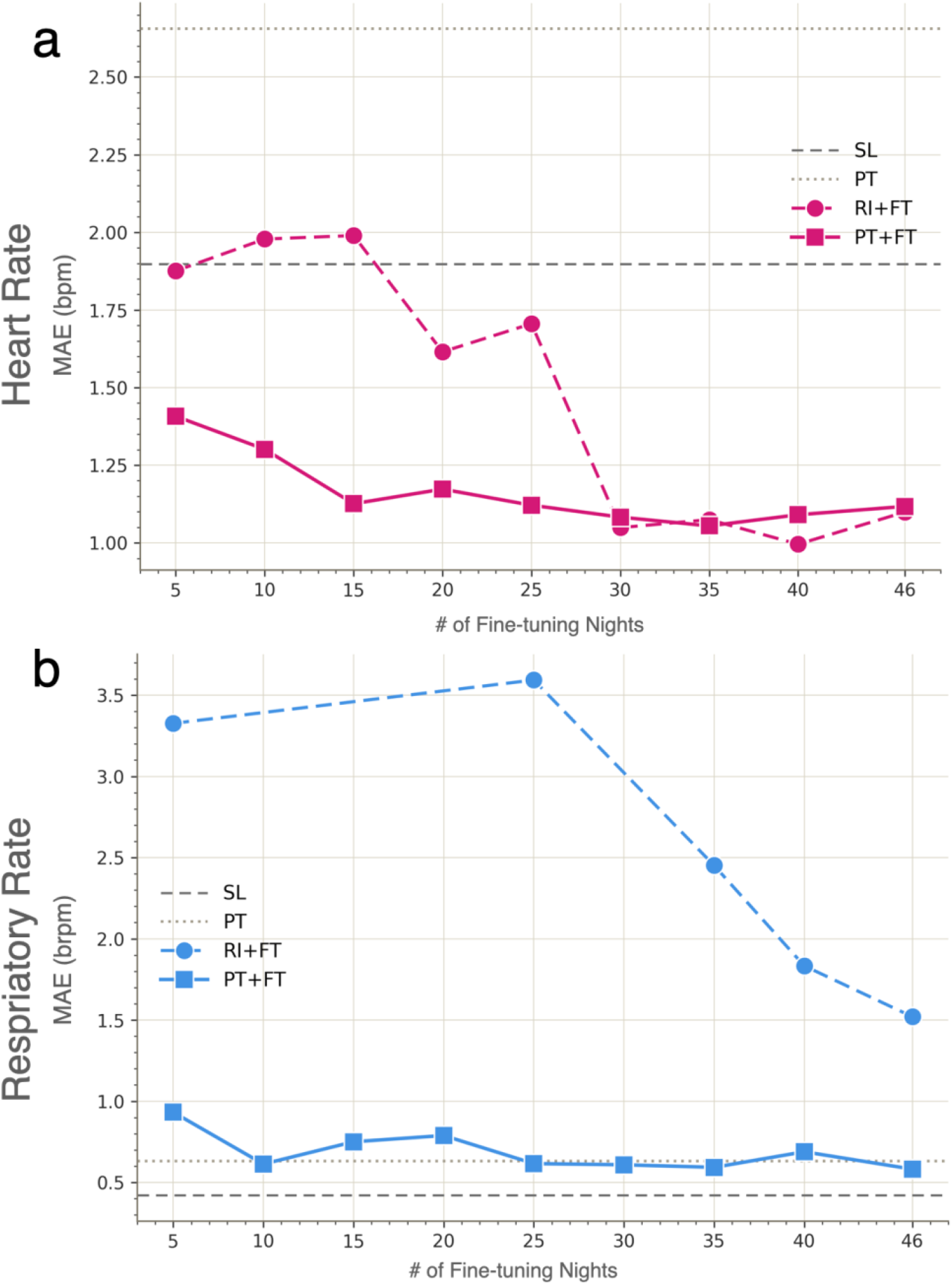
Pretraining improves sample efficiency during fine-tuning of bed-based sensor cardiopulmonary quantification models. For heart rate (HR) and respiratory rate (RR), pretrained (PT) models and randomly initialized (RI) models were fine-tuned (FT) on progressively fewer labeled data samples (from n=46 to n=5) and evaluated on the same n=60 held-out PSGs to avoid data leakage. They were compared to heuristic labeled data (SL - dashed line) and pretrained-only (PT - dotted line) models without fine tuning. Plots compare MAE for A) HR (n=20 models) and B) RR (n=20 models) as a function of amount of labeled data used for fine tuning. The RR RI+FT model produced no valid windows that pass quality gate rules at N = 10,15,20, and 30.

### Label efficiency during PSG-supervised fine-tuning

To test the hypothesis that pretraining can reduce the number of PSG-labeled nights required for fine-tuning, we trained PT+ FT and RI+FT models using progressively smaller subsets of 46-night fine-tuning data, down to as low as 5 nights. We used the same consistent 60-night --out test set for evaluation and comparison to avoid data leakage. For the smallest training sets, mimicking situations where labeled data is most limited, the pretrained PT+FT models markedly outperformed the randomly initialized RI+FT models. As the amount of labeled fine-tuning data increased, the benefits of pretraining waned until there was eventually no substantial performance difference between the approaches when evaluated on the 60 held-out datasets. For RR, the benefit of pretraining persisted across the full range of fine-tuning sample sizes examined (Fig. 3).

### Signal quality robustness

Waveform data varies in quality across time and between individuals. The ability to make high accuracy estimates on the maximum number of 1-minute windows indicates its robustness to signal quality. After performing model inference for RR and HR at each 1-minute window, we applied a quality filter (see methods). Whereas HR heuristic soft labels resulted in quality pass rate of 70%, the use of trained deep learning models all increased the pass rate to 90%, suggesting the benefit resulted from the flexibility of the trained model rather than whether we performed pretraining or fine-tuning. In comparison, the RR heuristic was already relatively robust to variation in signal quality, with a pass rate of 80%, and improved to 88% with the trained deep learning models (Tables 2–3).

## Discussion

Training and validation of longitudinal sensor data requires head-to-head ground truth labeled data that can be challenging and costly to collect. Far more abundant is real-world unlabeled data. Here, we show that a non-optimized peak-counting heuristic can be used to soft-label sensor waveforms to enable pretraining large real-world unlabeled datasets. This provided 2 major benefits. It required smaller amounts of head-to-head labeled data to create more performant models. It was also more robust to hyperparameter selection when tested across model sizes and learning rates. Although our study was conducted using non-contact bed sensors, the strategy should generalize to any longitudinal sensor for which large amounts of unlabeled real-world longitudinal data is available and labelled head-to-head ground truth data is scarce.

The resulting HR and RR mean absolute errors of 1.1 bpm and 0.6 brpm on 1-minute windows are comparable to performance reported in prior contactless validation studies of similar bed-sensor devices against polysomnography [13]. Because RR was evaluated against an algorithmic chest-RIP-derived reference rather than manual breath-by-breath annotations, RR performance should be interpreted as agreement with an independent PSG respiratory-effort signal.

This work is a specific case of a more general strategy in which pretraining a deep learning model on a large quantity of lower quality labels improves the sample efficiency of fine-tuning on high-quality labels, which are often hard to collect in large quantities [16, 17, 18]. We expect that this will be broadly useful for the sensor developer community who often have large amounts of real-world unlabeled data and comparatively limited amounts of high-quality head-to-head labelled data [9, 10]. This is particularly important for assembling FDA clearance datasets. Pretraining reduces the labeled data needed for algorithm development, which frees more of the limited head-to-head corpus for testing. Held-out test set sample sizes can then be driven by the need to adequately represent interpatient diversity rather than by training-set demands.

Our study has several limitations. We only examined one heuristic algorithm for soft labeling of RR and HR. Alternatives would likely have led to different degrees of improvement over random initialization. However, it is likely that the overall trends would remain the same. Consistent with our respiratory data, if the heuristic labels are exceptional, there will be little to gain from deep learning. On the other hand, if the heuristic is very poor and approach random, then the benefits of soft labels will wane and ultimately disappear. Even if soft labels are not used, there are other methods for label-free model pretraining that we did not explore. Such self-training strategies could also improve upon random initialization and could learn to capture the variation in the large amount of unlabeled real-world sensor data. Masked autoencoding is an example of such a strategy and would be worthwhile implementing and comparing in the future [19, 20]. Additionally, we only examined one model architecture and two hyperparameters. A further opportunity is that the model, although trained to predict a specific rate, likely learns representations that capture additional properties of the underlying signal. Such pretrained features have been shown to transfer to tasks beyond the original training objective [21, 22], suggesting that the same encoder could be repurposed for related downstream tasks such as sleep staging, arousal detection, or apnea identification with minimal additional supervision.

Our analysis revealed two benefits of pretraining. First, it reduced the amount of PSG-labeled data required to obtain performant models, with the strongest HR benefit at low sample size and a persistent RR benefit across the full sample-size range. Second, it reduced sensitivity to architecture size and learning-rate selection. The final accuracy benefit was task-dependent: with 46 labeled nights, HR RI+FT and HR PT+FT performed similarly, whereas RR PT+FT substantially outperformed RR RI+FT. One likely explanation is that the RR heuristic was substantially more accurate than the HR heuristic. Such pretraining on a stronger soft-label teacher provides a better starting point that fine-tuning on 46 nights of head-to-head data cannot fully replace. Taken together, these results suggest that domain heuristics - widely available in mature sensor pipelines - can serve as a practical bridge between abundant unlabeled real-world data and scarce gold-standard labels for training validated quantification models.

## Methods

### Datasets

Sensor data was collected with a thin, flexible ferroelectret mechanical transducer positioned beneath the mattress (referred to as ‘bed sensor’). The device captures the mechanical forces of cardiac contraction, respiration, and gross body movement, and its raw output is decomposed into a high-frequency band (ballistocardiogram) and a low-frequency band (respiratory waveform). These signals serve as input to all quantification models described below. Two non-overlapping datasets from Nightingale Labs were used. Dataset #1 is a real-world corpus of 50,848 overnight recordings from 395 adult subjects sleeping at home, recorded longitudinally, with a median of *∼*100 nights per subject and no clinical reference signal. It was used exclusively for pretraining.

Dataset #2 is a corpus of 106 overnight recordings, each from a distinct subject, with simultaneous clinical-grade polysomnography (PSG) and bed sensor recording. From the PSG signals, Electrocardiography provided HR ground truth and chest respiratory inductance plethysmography (chest-RIP) provided RR ground truth. Dataset #2 was partitioned at the subject level into a 46-night fine-tuning pool and a 60-night held-out test set; the test set was sequestered from all training and hyperparameter-selection decisions. The two datasets share no subjects.

### Heuristic algorithms

Two deterministic heuristic algorithms estimated heart rate and respiratory rate, serving both to generate soft labels (SL) for pretraining on Dataset #1 and as baseline references evaluated directly on the held-out test set. Both share a common core structure: the relevant channel is bandpass-filtered and divided into fixed-length windows (60 seconds for the low-band RR channel, 30 seconds for the high-band HR channel). Within each window, an adaptive-midline scheme selects the amplitude threshold yielding the most regularly spaced peaks. After discarding physiologically implausible peak-to-peak intervals (outside 1–10 s for RR, and 0.3–2.0 s for HR), the window’s rate is calculated as the reciprocal of the median retained interval. The respiratory rate heuristic applies this logic directly to the low-band channel. Conversely, the heart rate heuristic operates on the high-band channel and first converts the bipolar ballistocardiogram waveform into a unipolar envelope, isolating heartbeats as distinct bumps. To prevent slow amplitude drifts from causing missed peaks, the HR candidate thresholds follow a moving mean of this envelope rather than flat lines.

### Neural model and preprocessing

We used a single-task neural network architecture, trained separately for heart rate (HR) and respiratory rate (RR). The model uses one-dimensional (1D) convolutions to extract local signal features that feed into a bidirectional Gated Recurrent Unit (GRU) to encode temporal patterns. This output is mean-pooled across time and passed through a Multi-Layer Perceptron (MLP) to predict a single scalar rate per window. We evaluated four size variants of this architecture, spanning 14 .6K to 902K parameters. Every model processes a 3,000-sample input window, corresponding to 30 seconds of high-band data for HR or 60 seconds of low band data for RR. Before entering the model, signals undergo task-specific preprocessing: HR inputs are bandpass filtered, rectified, and smoothed to emphasize individual heartbeats, whereas RR inputs are only bandpass filtered. Finally, to handle natural fluctuations in signal strength, each window is independently normalized using a robust z-score based solely on its own median and interquartile range.

### Stage 1: heuristic-supervised pretraining

During this stage, models were pretrained on Dataset #1 to predict the heuristic SL. The dataset was split at the subject level (70/15/15 train/validation/test), and only the 277-subject training split was used to construct the pretraining pool. Within each night, we generated candidate windows (30 s non-overlapping for HR; 60 s with a 30 s stride for RR) and passed them through the corresponding heuristic soft labelling algorithm. Windows were retained only if the generated SL fell within physiological limits (25–220 BPM for HR; 2–80 brpm for RR), effectively discarding segments of poor signal quality.

Because the bed-sensor’s cardiac signal is mechanically weaker than the respiratory signal, the HR SL are inherently noisier. To account for this, HR windows underwent an additional stability filter, retaining only those that fell within a contiguous 5-minute span of stable heart rate. This yielded final pretraining pools of 15.3 million HR windows and 42.9 million RR windows. Each of the four architecture widths was trained independently for both tasks (eight runs total). Models were trained for 10 epochs using Huber loss (*β* = 1.0) and the AdamW optimizer (learning rate 5 *×* 10^*−*4^, batch size 256). For computational tractability, each epoch sampled a random subset of 2 million windows. The final epoch-10 checkpoints were saved for downstream fine-tuning and evaluation on the held-out test set.

### Stage 2: PSG-supervised fine-tuning

In this stage, models were fine-tuned on the 46-night fine-tuning pool from Dataset #2 using PSG-derived ground-truth labels. We trained models under two initialization conditions: PT+FT, which initialized weights from the matching Stage-1 checkpoint, and RI+FT, which trained the same architecture with random initialization. For PT+FT models we applied L2-Starting Point (L2-SP; *λ* = 1 *×* 10^*−*5^) regularization to anchor the new weights near their pretrained values. To maximize the number of training examples, this stage used overlapping windows (a 1-second sliding stride for HR and a 10-second stride for RR), exposing the model to many shifted views of each labeled epoch.

HR ground-truth labels were calculated from the simultaneous ECG channel using R-peak detection. RR ground-truth labels were derived by applying the respiratory heuristic separately to the chest-RIP and abdominal-RIP channels, meaning a single sensor window could contribute to two separate training examples. The 46-night pool was split at the subject level, reserving a small validation subset (7 nights for HR, 3 nights for RR) to monitor for early stopping. For each task and initialization condition, we searched a grid combining the four architecture widths with four candidate learning rates, ultimately retaining the model configuration that achieved the lowest validation loss.

### Test-set inference and ground truth

We evaluated the final models on the 60 held-out test nights. First, the model analyzed each recording by sliding forward one second at a time, producing a new prediction every second. To aggregate these second by-second predictions, we grouped them into non-overlapping 1-minute bins by taking the median value for each minute. Next, we established an independent clinical “ground truth” for each minute to grade the model against. For heart rate, we used the simultaneous ECG channel: we detected individual heartbeats (R-peaks via BioSPPy and NeuroKit2 [23, 24]), discarded invalid intervals, and calculated the reference rate by averaging the remaining clean intervals. For respiratory rate, reference labels were derived from the clinical chest-RIP signal acquired during polysomnography by applying the RR heuristic to overlapping 60-second windows and aggregating the values. Because the RIP signal directly measures respiratory effort and has substantially higher respiratory signal to-noise than the bed-sensor waveform, it served as the PSG-derived reference for RR evaluation.

### Quality gate

Performance metrics on the test set were only evaluated in the windows that passed a six-rule quality gate Table 1. This gate evaluates the stability of the model’s predictions alongside the integrity of the PSG reference signal. The final fraction of retained data for each model is reported as its bin-pass percentage in Table 2. Optimal thresholds for the rules were selected using grid search in the training data set. These thresholds were applied as it is to the test set inference without any further optimization.

**Table 1.** The 6 rules that every 1-minute window must satisfy in the held-out 60-night evaluation set. Rules R1 to R4 are based on the outputs of the model being evaluated. Rules R5 and R6 evaluate the quality of the ground truth label derived from PSG signals.

| Rule | Criteria / Threshold | Rationale |
| --- | --- | --- |
| R1 Variance Lower Bound | Within-bin variance of dense (1-s stride) predictions must be $\geq 0.005$ (HR) / $0.0005$ (RR). | Rejects flat prediction streams, which indicate either a featureless sensor input or model collapse. |
| R2 Constant-Fallback Removal | Predictions within $\pm 5 \times 10^{-4}$ of the model's validation-derived modal output are excluded | Ensures metrics reflect active signal tracking rather than the model falling back on a default guess. |
| R3 Variance Upper Bound | Within-bin variance of dense predictions must be $\leq 30$ (HR) / $5$ (RR). | Rejects wildly oscillating prediction streams typically caused by motion artifacts. |
| R4 Streak Extension | Rejects bins immediately adjacent to a streak of three or more consecutive R1 failures. | Removes transitional data contaminated by the boundary of an event (e.g., getting off the bed). |
| R5 Ground-Truth Plausibility | Reference rate cannot deviate by $> 10$ BPM (HR) / $2$ brpm (RR) from its local 5-bin centered median. | Catches isolated reference-channel computation errors (R-peak miscounts for HR; chest-belt detection artefacts for RR). |
| R6 Reference Signal Integrity | Per-bin raw variance of the reference channel must be $\geq 5\%$ of the night's median variance (ECG channel for HR / chest belt for RR). | Catches reference sensor detachment (ECG lead-off for HR; chest-belt fall-off for RR), which invalidates the ground-truth rate. |

**Table 2.** Comparison of pretrained and fine-tuned models for bed-based sensor cardiopulmonary quantification. The table compares 8 models of which 4 estimate heart rate (HR) and 4 estimate respiratory rate (RR). For each metric we compare heuristic-labeled data (SL), pretrained without fine-tuning (PT), random initialization with fine-tuning (RI+FT), and pretrained with fine-tuning (PT+FT). Columns reflect the following error metrics - MAE: Mean absolute error, MedAE: Median Absolute Error, RMSE: Root Mean Square Error, SD: Standard Deviation of Error curve. N bins refers to the number and percentage of 1 minute epochs that pass the quality filters (see methods).

| method | # params | N bins | Pass % | MAE | MedAE | RMSE | Bias | SD | unit |
| --- | --- | --- | --- | --- | --- | --- | --- | --- | --- |
| HR SL | - | 18311 | 68.5% | 1.9 | 0.71 | 5.35 | 0.64 | 5.31 | bpm |
| HR PT | 226k | 23643 | 88.4% | 2.66 | 1.25 | 6.01 | 0.37 | 6 | bpm |
| HR RI+FT | 902k | 24063 | 90% | 1.11 | 0.64 | 2.21 | 0.1 | 2.21 | bpm |
| HR PT+FT | 226k | 23856 | 89.2% | 1.07 | 0.6 | 2.21 | 0.18 | 2.2 | bpm |
| RR SL | - | 21633 | 80.9% | 0.42 | 0.2 | 0.88 | 0.04 | 0.88 | brpm |
| RR PT | 226k | 23285 | 87% | 0.63 | 0.27 | 1.4 | 0.25 | 1.38 | brpm |
| RR RI+FT | 226k | 24177 | 90.4% | 1.42 | 0.9 | 2.15 | -0.57 | 2.07 | brpm |
| RR PT+FT | 226k | 23760 | 88.8% | 0.59 | 0.32 | 1.15 | 0.21 | 1.13 | brpm |

**Table 3.** Comparison of pretrained and fine-tuned models for bed-based sensor cardiopulmonary quantification for a common set of epochs. The table compares 8 models of which 4 estimate heart rate (HR) and 4 estimate respiratory rate (RR). For each metric we compare heuristic-labeled data (SL), pretrained without fine-tuning (PT), random initialization with fine-tuning (RI+FT), and pretrained with fine-tuning (PT+FT). Columns reflect the following error metrics - MAE: Mean absolute error, MedAE: Median Absolute Error, RMSE: Root Mean Square Error, SD: Standard Deviation of Error curve. N bins refers to the number and percentage of 1 minute epochs that pass the quality filters (see methods).

| method | #params | N bins | Pass % | MAE | MedAE | RMSE | Bias | SD | unit |
| --- | --- | --- | --- | --- | --- | --- | --- | --- | --- |
| HR SL | - | 17569 | 65.7% | 1.54 | 0.68 | 3.8 | 0.4 | 3.78 | bpm |
| HR PT | 226k | 17569 | 65.7% | 1.61 | 1.08 | 3.45 | -0.22 | 3.45 | bpm |
| HR RI+FT | 902k | 17569 | 65.7% | 0.9 | 0.58 | 1.71 | 0.18 | 1.7 | bpm |
| HR PT+FT | 226k | 17569 | 65.7% | 0.86 | 0.54 | 1.69 | 0.29 | 1.67 | bpm |
| RR SL | - | 21305 | 79.6% | 0.41 | 0.2 | 0.86 | 0.04 | 0.86 | brpm |
| RR PT | 226k | 21305 | 79.6% | 0.47 | 0.25 | 0.94 | 0.21 | 0.92 | brpm |
| RR RI+FT | 226k | 21305 | 79.6% | 1.19 | 0.81 | 1.76 | -0.53 | 1.67 | brpm |
| RR PT+FT | 226k | 21305 | 79.6% | 0.46 | 0.29 | 0.8 | 0.2 | 0.77 | brpm |

### Data-efficiency sweep

To assess how performance scales with PSG-labeled data, the 46-night fine-tuning pool was shuffled once and progressively restricted to nested subsets of N *∈* {5, 10, 15, 20, 25, 30, 35, 40, 46} nights. The subject-level train/validation split was re-applied within each subset. For each task and initialization condition, a single Stage-2 fine-tuning run was performed using the optimal architecture width and learning rate identified from the full 46-night grid search. Performance (Mean Absolute Error) was then evaluated on the unchanged 60-night test set.

### Statistical analysis

Performance metrics were computed across all 1-minute bins that passed the quality gate, pooled across the 60 test nights. Per-bin error was defined as the model prediction minus the ground truth. We calculated Mean Absolute Error (MAE), median absolute error (MedAE), root-mean-square error (RMSE), bias, standard deviation (SD), and Bland-Altman 95% limits of agreement (bias *±* 1.96 *×* SD) in the rate’s natural units (BPM for HR, brpm for RR). To isolate the comparison from differences in data retention across models, a matched-epoch variant of these metrics was also computed by restricting the evaluation to the exact intersection of bins that passed the quality gate across all four evaluated methods (SL, PT, RI+FT, PT+FT).

## Data Availability

The datasets analyzed during the current study are not publicly available due to participant privacy and data-use restrictions but may be available from the corresponding author on reasonable request and subject to appropriate institutional approvals and data-use agreements. Code used for preprocessing, model training, evaluation, and figure generation may be made available from the corresponding author on reasonable request.

## Ethics approval and consent to participate

The study was approved by the University of California San Diego Institutional Review Board (IRB protocol #171480; Principal Investigator: King), and all participants provided written informed consent. All procedures were performed in accordance with relevant guidelines and regulations. Informed consent was obtained from all participants or their legally authorized representatives.

## Acknowledgements

We thank the UCSD Research Sleep Lab for their assistance with head-to-head PSG collection. We thank Nightingale Lab for use of sensors and sensor data. This work was funded by the American Heart Association (AHA) Predoctoral Fellowship 26PRE1560767 (R.P.V.). NIH NHLBI T32 HL007444 Training Program on Cardiovascular Physiology and Pharmacology Program (D.T.B.); NIH R33 HL168785, R43HL157291, R43AI177239, DOD HT94252410105, and Wellcome Leap In Utero Program (K.R.K.). This publication was supported in part by the Altman Clinical and Translational Research Institute at the University of California, San Diego, funded by the National Center for Advancing Translational Sciences, National Institutes of Health, Award Number UM1TR005449. The content is solely the responsibility of the authors and does not necessarily represent the official views of the NIH.

## Author contributions statement

K.G. developed the analysis pipeline, trained and evaluated the models, analyzed the results, prepared the figures and tables, and drafted the manuscript. N.H. contributed to study design, heuristic algorithm development, and manuscript revision. R.P.V. contributed to analysis, and manuscript revision. P.D., R.L.O., and J.O. contributed clinical and sleep-medicine expertise, interpretation of polysomnography-derived measures, and manuscript revision. K.R.K. conceived and supervised the study, contributed to interpretation of results, and revised the manuscript. All authors reviewed and approved the final manuscript.

